# Spatio-temporal analysis of spotted fever cases reported to a tertiary care hospital in Southern India

**DOI:** 10.64898/2026.04.07.26350285

**Authors:** Teena Mariam Thomas, Solomon D’Cruz, Susmitha Karunasree Perumalla, Karthik Gunasekaran, John Antony Jude Prakash

## Abstract

**Background:** Spotted fever is caused by spotted fever group rickettsiae (SFGR) belonging to the genus Rickettsia. Transmission to humans is primarily via the bite of infected ticks. Being a vector-borne disease, the occurrence of spotted fever is related to factors that allow the vector to thrive. This spatio-temporal analysis gives an insight into the distribution of cases and correlation with seasonality.

**Methodology:** A suspected AFI patient was considered spotted fever positive if either serology (ELISA/IFA) or molecular assay (Nested PCR/qPCR) was tested positive. Demographic data of confirmed cases were included for the analysis.

**Results:** In the 18-year dataset, a total of 2153 suspected patients were tested for spotted fever, of which 516 (24%) were positive. On spatio-temporal analysis, Vellore district reported 39.9% of cases, Chittoor 38.8%, Tirupattur 12.5%, Ranipet 4.5%, and Tiruvannamalai 4.3%. Maximum spotted fever cases were reported between the months of September to March, with a peak in January. Children below 10 years and housewives were at risk of spotted fever.

**Conclusion:** The findings of this retrospective analysis highlight the importance of considering spotted fever group rickettsioses in patients presenting with acute undifferentiated febrile illness, particularly children aged <10 years, from areas with higher spatial clustering, during or following the monsoon season.

## Introduction

Acute Undifferentiated Febrile Illness (AUFI) is defined as a febrile illness of ≤15 days duration without an identifiable localising cause at presentation (1). Among these, rickettsial infections like scrub typhus and spotted fever are one of the major causes of AUFI (2).

Spotted fever is caused by Spotted fever group rickettsiae (SFGR) belonging to the genus Rickettsia (3). These are obligate intracellular gram-negative bacteria that are maintained in nature through complex transmission cycles involving arthropod vectors and vertebrate reservoir hosts (4). *Rickettsia conorii* is the causative agent for Mediterranean Spotted Fever (MSF) in several parts of Europe, Africa and Asia (5). Transmission to humans is primarily via the bite of infected ticks. The ticks are hematophagous arthropods categorised as Ixodidae (hard ticks) and the Argasidae (soft ticks) with different morphological and feeding habits (6). The presence of bacteria in the salivary glands of ticks is of significant importance, as it facilitates the transmission of Rickettsia during feeding (7). In India, Rhipicephalus sanguineus is considered the principal vector of Indian tick typhus; humans are incidental (dead□end) hosts (8). Domestic animals such as dogs serve as important hosts, facilitating human exposure in rural and peri-domestic settings (9).

The clinical features of spotted fever consist of fever with rash, most commonly maculopapular, characteristically involving the palms and soles (10). However, the clinical spectrum is variable, and the absence of rash in a subset of patients can make diagnosis challenging. Laboratory confirmation of spotted fever relies on serology assays like Enzyme Linked Immunoassay (ELISA), Immunofluorescence antibody (IFA) and molecular assays like PCR (6). The drug of choice is doxycycline or a combination of doxycycline and azithromycin is also effective for spotted fever (11).

Being a vector-borne disease, the occurrence of spotted fever is related to factors that allow the vector to thrive. Environmental factors such as temperature, humidity, and vegetation play a crucial role in determining tick survival and distribution (12). Rickettsial infections have been reported to exhibit seasonal trends with regional variations (6). This study focuses on the spatial and temporal distribution of spotted fever cases, diagnosed at a tertiary care centre in Vellore over a period of 18years (2007 – 2024).

## Materials & Methods

### Methodology

From January 2007 to December 2024 (18 years data), 2153 patients with a clinical suspicion of spotted fever were tested by serology and molecular assays in the Immunology laboratories (an ISO15189:2012 accredited lab), Department of Clinical Microbiology, Christian Medical College, Vellore. The demographic data, date of testing and results were obtained from the electronic medical records of the institution.

### Serology

From January 2007 – September 2009, IgM ELISA for spotted fever group rickettsiae (SFGR) was performed using a commercial kit (PanBio Ltd, Brisbane, Australia) and a value of ≥16 units was considered positive. The Rickettsia Screen IFA IgM Antibody Kit (Fuller Laboratories, Fullerton, CA, USA) was used from October 2009 to May 2015, IgM IFA titre of 1:64 was considered positive. Since June 2015, Rickettsia conorii IgM ELISA (Vircell, Granada, Spain) with an OD value of ≥ 1 is considered as positive (Serum dilution 1:25 as per manufacturer instructions) for spotted fever.

### Molecular

From 2007 – 2008, nested PCR to amplify spotted fever genes was performed on 58 skin biopsies and amplicon size (532bp) was confirmed by gel electrophoresis (10). Since 2018, a real-time PCR (qPCR), which amplifies a 100bp segment of the spotted fever-specific ompA gene, has been performed for 193 samples as per the previously published protocol (1). For the real-time PCR for the OmpA gene, a Ct value of ≤ 38 is considered positive for spotted fever.

### Positive Results

A patient suspected with spotted fever is considered positive if IgM antibodies for spotted fever were detected by serology and/or the OmpA gene was identified using PCR (nested PCR or quantitative PCR).

### Statistical analysis

The demographic data and the results were entered to Microsoft Excel version 2015 spreadsheets, and analysis was performed using IBM SPSS Statistics for Windows, Version 21.0 (IBM Corp, Armonk, NY, USA). The continuous variables were expressed in mean and standard deviation (SD) for normal distribution. The categorical variables were expressed in frequencies and percentages. The association between two categorical variables was analysed using chi-square, and logistic regression was used to adjust for confounders.

### Study Area

The study districts for spatio-temporal analysis included Vellore, Ranipet, Tirupattur, Tiruvannamalai, and Chittoor. These are the major catchment areas and are characterised by a mix of urban, peri-urban, and predominantly rural settlements. Geographically, they lie within the eastern part of peninsular India and are marked by undulating terrain interspersed with hilly ranges, including the Eastern Ghats. The climate is tropical, with hot summers, moderate winters, and rainfall primarily influenced by both the southwest and the northeast monsoon.

### Spatiotemporal analysis

The QGIS 3.34.3 software was used for spatial analysis at the sub-district (taluk) level. The shape files were downloaded from the Survey of India website (https://surveyofindia.gov.in/). A seasonal trend analysis was undertaken to depict the peak month of spotted fever cases using R software version 4.5.2. A negative binomial regression model was used to assess the association between monthly variation and the number of spotted fever cases, accounting for overdispersion in the data.

### Ethical Statement

The approval to conduct this study was obtained from Institutional Review Board (IRB) and Ethics Committee (EC) of the Christian Medical College (CMC), Vellore, IRB Min no. 0624131 dated 12.06.2024. The required data had been collected as part of the standard of care for diagnosis of Spotted fever, informed consent waiver was granted by the IRB and EC of CMC, Vellore. No human subjects were directly involved in the study.

## Results

From 2007 – 2024, among the 2153 patient samples tested, 465 & 16 samples were exclusively positive by serology and OmpA PCR, respectively, whereas both serology and PCR positives were 35. Effectively, spotted fever positive cases were 516 with an overall positivity rate of 24% (Range: 5 - 39%), refer supplementary data (Suppl Fig.1&2).

### Demographic & Risk Factor Analysis

The male-to-female ratio of samples tested is 27:23. The mean age of the participants was 30 years, with SD of 21.1 years. The demographic characteristics of study participants, including age, gender, OP/IP, and occupation are given in Table 1.

**Table 1.** Demographic Characteristics.

| **n=2153** | **Total Tested** | **SF Negatives** | **SF Positives** | **Positivity Rate\*** |
| --- | --- | --- | --- | --- |
| **Age group** |  |  |  |  |
| 0 - 10 years | 520 | 299 | 221 | 42.5% |
| 11 – 20 years | 261 | 231 | 50 | 19.2% |
| 21 - 30 years | 387 | 320 | 67 | 17.3% |
| 31 - 40 years | 296 | 249 | 47 | 15.9% |
| 41 - 50 years | 241 | 195 | 46 | 19.1% |
| 51 - 60 years | 228 | 179 | 49 | 21.5% |
| >60 years | 200 | 164 | 36 | 18.0% |
| **Gender** |  |  |  |  |
| Male | 1168 | 890 | 278 | 23.8% |
| Female | 985 | 747 | 238 | 24.2% |
| **OP/IP** |  |  |  |  |
| IP | 1570 | 1257 | 313 | 19.9% |
| OP | 583 | 380 | 203 | 34.8% |
| **Occupation** |  |  |  |  |
| Agriculture | 92 | 76 | 16 | 17.4% |
| Daily wage Worker | 154 | 125 | 31 | 20.1% |
| House wife | 357 | 279 | 78 | 21.8% |
| Students | 854 | 581 | 273 | 32.0% |
| Unemployed | 368 | 298 | 70 | 19.0% |
| Others | 328 | 280 | 48 | 14.6% |

\*Positivity rate: Number of positives/Samples tested for each demographic category. Hence, the percentage values are not additive.

Table 2 depicts the odds ratio (OR) and adjusted odds ratio (AOR) by logistic regression assessing the association of gender, age, and occupation with spotted fever.

**Table 2.** Association of gender, age, and occupation with spotted fever.

| Variable |  | SF Pos | SF Neg | OR (95%CI) | p-Value | AOR (95% CI) | p-Value |
| --- | --- | --- | --- | --- | --- | --- | --- |
| Gender | Female | 238 | 747 | 0.98 (0.80-1.20) | 0.845 | 1.016 (0.79-1.29) | 0.899 |
|  | Male | 278 | 890 |  |  |  |  |
| Age | Above 60 yrs | Reference |  |  |  |  |  |
|  | 0 – 10 years | 221 | 299 | 3.37 (2.26-5.03) | <0.001 | 5.03 (2.69-9.40) | <0.001 |
|  | 11 – 20 years | 50 | 231 | 0.99 (0.61-1.58) | 0.954 | 1.37(0.74-2.51) | 0.315 |
|  | 21 – 30 years | 67 | 320 | 0.95 (0.610-1.491) | 0.836 | 1.06 (0.66-1.67) | 0.815 |
|  | 31 – 40 years | 47 | 249 | 0.86 (0.53-1.39) | 0.535 | 0.90 (0.55-1.45) | 0.659 |
|  | 41 – 50 years | 46 | 195 | 1.07 (0.66-1.74) | 0.770 | 1.11 (0.67-1.80) | 0.682 |
|  | 51 – 60 years | 49 | 179 | 1.25 (0.77-2.01) | 0.367 | 1.28 (0.78-2.07) | 0.319 |
| Occupation | Others | Reference |  |  |  |  |  |
|  | Agriculture | 16 | 76 | 1.23 (0.66-2.28) | 0.516 | 1.25 (0.67-2.34) | 0.478 |
|  | Daily wage Worker | 31 | 123 | 1.47 (0.89-2.42) | 0.130 | 1.49 (0.90-2.46) | 0.116 |
|  | Housewife | 78 | 279 | 1.63 (1.09-2.42) | 0.015 | 1.66 (1.07-2.57) | 0.020 |
|  | Unemployed | 70 | 298 | 1.37 (0.91-2.04) | 0.125 | 1.37 (0.91-2.06) | 0.128 |
|  | Students | 273 | 581 | 2.74 (1.95-3.84) | <0.001 | 0.93 (0.53-1.61) | 0.780 |

The analysis showed children aged 0 –10 years at significantly higher risk, with an AOR of 5.03 (95% CI: 2.69–9.40; p < 0.001) compared to the reference group. Occupation-wise, housewives had a significantly increased risk (AOR: 1.66; 95% CI: 1.07–2.57; p = 0.02).

### Spatio-temporal analysis

The cases tested and the positivity rate throughout the study period (2007-2024) is given in Supplementary Fig 1 & 2. Among the 516 spotted fever positive cases, 441 cases were reported from Vellore and the 4 adjoining districts, Chittoor, Tirupattur, Ranipet, and Tiruvannamalai (Fig. 1).

**Fig 1:**
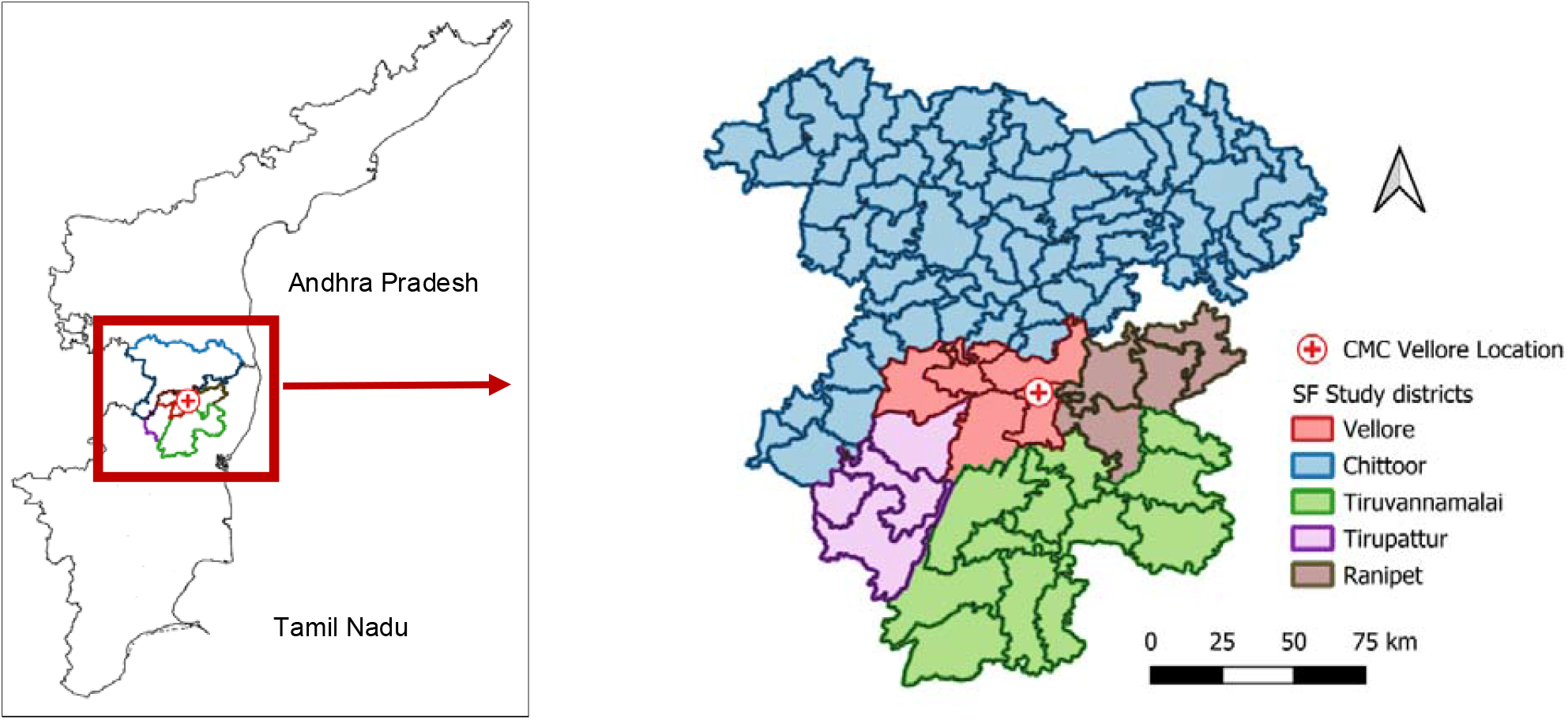
The districts with the taluk-wise depiction from where the 441 cases were reported

Among these 5 districts, Vellore district reported 39.9% of cases, Chittoor 38.8%, Tirupattur 12.5%, Ranipet 4.5%, and Tiruvannamalai 4.3% (refer Suppl Table.1). The spatio-temporal analysis of case occurrence in the 5 districts at the taluk level is given in Fig.2.a-d. List of taluks reporting spotted fever cases is as given in Suppl Table 2. Taluk-level spatial depiction was used to provide a finer resolution of case distribution, enabling the identification of localised clustering patterns.

**Figure 2:**
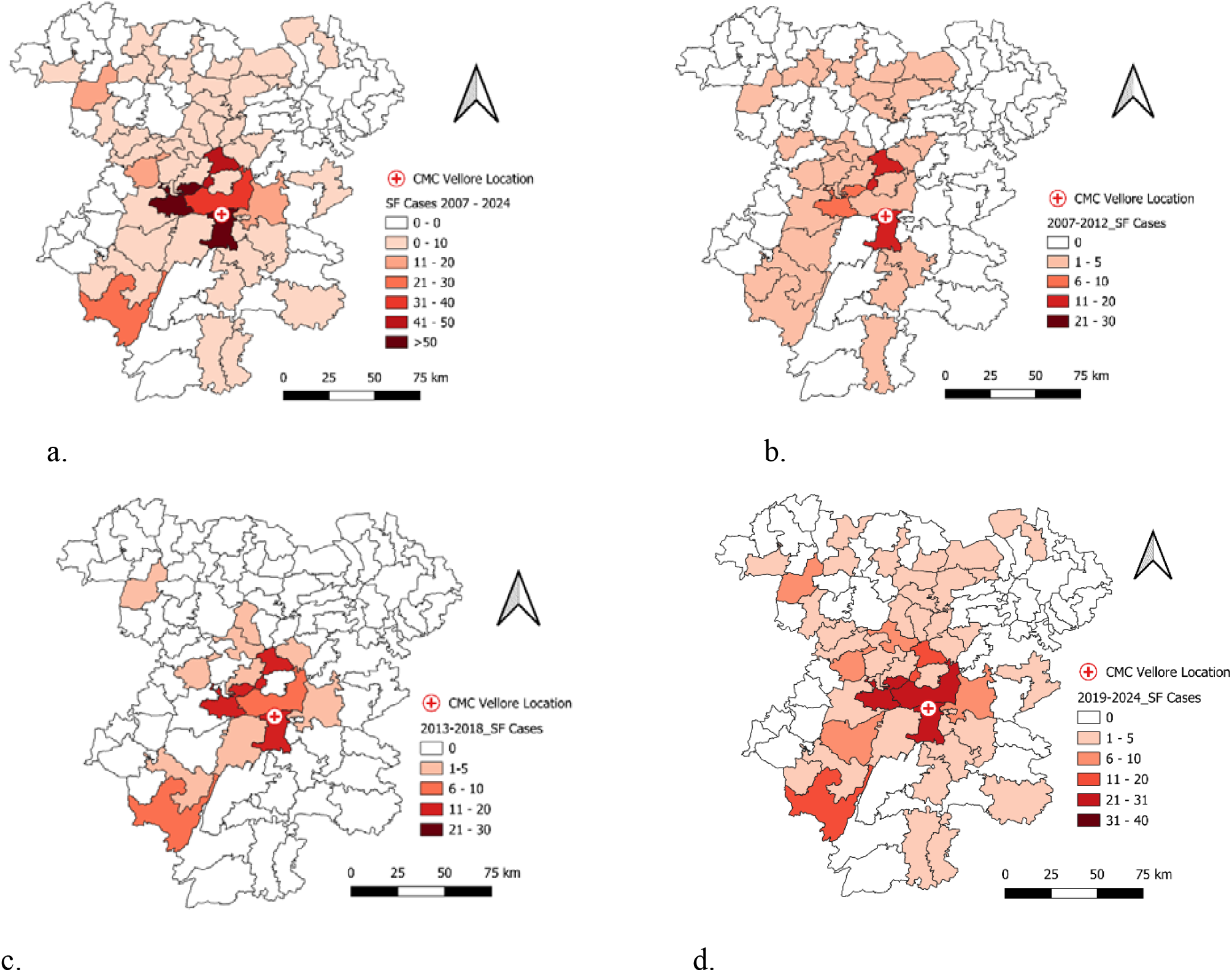
Spatio-temporal analysis of spotted fever cases in the 5 districts.

### Seasonal trend analysis

The 18-year seasonal trend analysis demonstrates cases occur throughout the year the gradual increase from September, peaking in January, followed by a declining trend in March (Fig 3). On negative binomial regression analysis, when compared to the month of July, there has been a gradual increase in the IRR from September, and attains the strongest association in January (IRR: 22.8, 95% CI (10.02 – 51.76), p <0.001). Thereafter, it starts decreasing, reaching a low level in March, and it is statistically significant (Refer Table 3).

**Fig.3:**
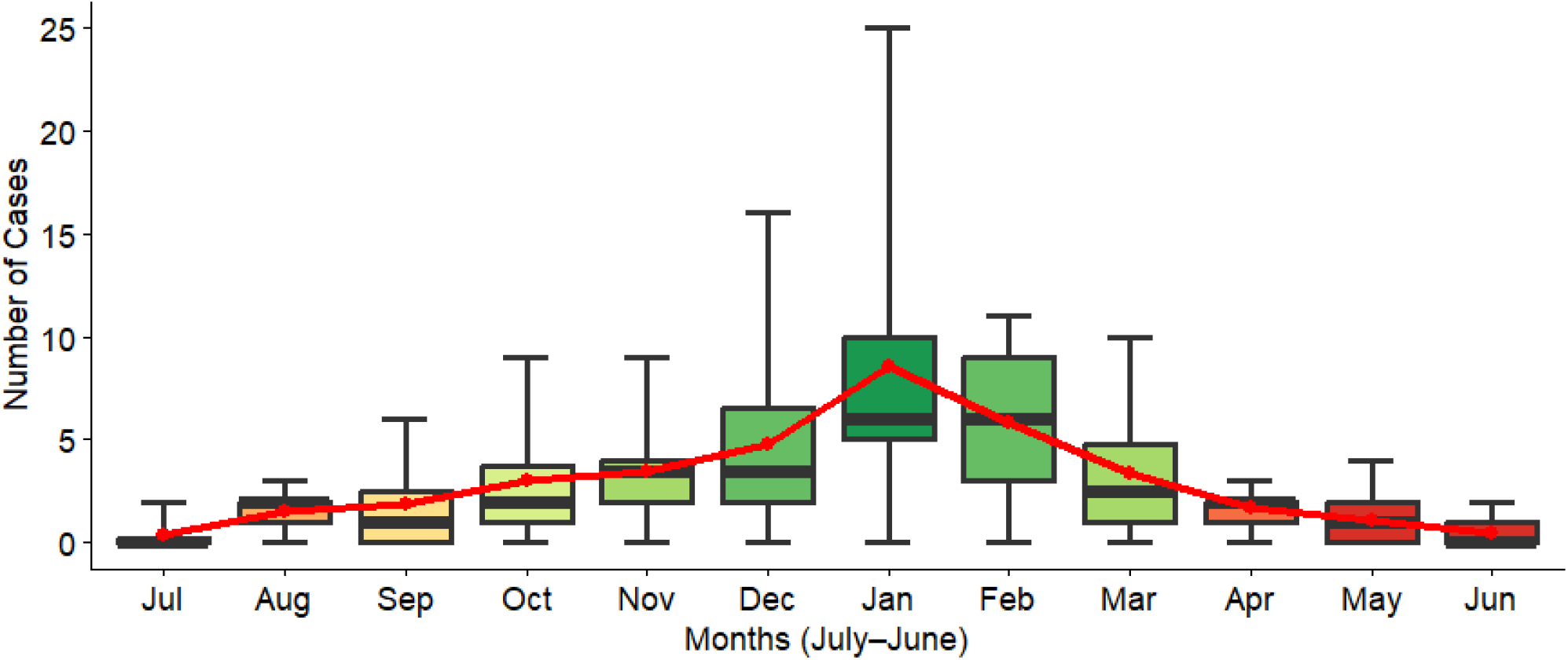
Month-wise distribution of spotted fever cases (2007-2024)

**Table 3.** Negative binomial regression analysis of monthly SF Cases (2007–2024)

| Months | IRR* | 95% CI | p-Value |
| --- | --- | --- | --- |
| July | Reference |  |  |
| August | 4 | 1.42 – 11.22 | 0.008 |
| September | 4.98 | 1.86 – 13.29 | 0.001 |
| October | 8 | 3.4 – 18.81 | <0.001 |
| November | 9.17 | 3.95 – 21.29 | <0.001 |
| December | 12.67 | 5.52 – 29.08 | <0.001 |
| **January** | **22.77** | **10.02 – 51.76** | **<0.001** |
| February | 15.64 | 6.84 – 35.76 | <0.001 |
| March | 8.95 | 3.83 – 20.93 | <0.001 |
| April | 4.51 | 1.64 – 12.37 | 0.003 |
| May | 2.84 | 1.01 – 7.99 | 0.047 |
| June | 1.24 | 0.38 – 4.02 | 0.714 |

\*IRR: Incidence Rate Ratio.

## Discussion

Spotted fever group rickettsiosis are emerging or re-emerging tick-borne diseases in humans (1). In this 18-year data analysis (retrospective), there were 516 spotted fever cases. The commonest age-group affected were children less than 10 years (43% of reported cases). Similar findings have been noted in Portugal (13) and from Mexico (14), whereas spotted fever has been found to be more common (≈ 85%) in individuals ≥15 years in Spain (15) and Italy (16). This variation may reflect differences in exposure patterns, with children in endemic rural settings having greater peridomestic exposure to tick-infested environments, whereas in adults, occupational exposure is more likely (17). Also, a serosurvey on rickettsial infection showed that the common occupation of patients was farmers (37%), followed by housewives (35%) (18). This correlates with findings of this study where housewives were found to be at a higher risk.

For spatio-temporal analysis, the study period was divided into three phases (2007–2012, 2013–2018, and 2019–2024) based on case occurrence. The first phase represented a low-incidence period (2007-2012), the second a transitional phase (2013-2018) with a gradual rise in cases, and the third a high-incidence phase (2019-2024). The observed temporal increase in cases may reflect the evolving clinical recognition and advancements in diagnostic practices over time. A study conducted at our centre during 2007–2008 contributed to the initial identification of spotted fever in the region (10).

Subsequently, a prospective study conducted among acute febrile cases in 2019 (1) with the inclusion of ELISA and real-time PCR likely enhanced clinical suspicion and increased testing thereafter.

Collectively, among the 516 spotted fever cases, 441 were reported from the 5 catchment districts (Vellore, Chittoor, Tirupattur, Ranipet, and Tiruvannamalai), which contribute the maximum numbers of acute febrile illness cases reporting to our centre. A taluk-level spatial analysis was performed to achieve a more granular understanding of case distribution, enabling the detection of localised clustering patterns that may not be apparent at the district level.

The higher burden of spotted fever cases is likely multifactorial, involving ecological suitability, land-use patterns and health-care access (12). The known risk factors for spotted fever include outdoor exposure to ticks, agricultural work and contact with domestic animals. These are more prevalent in rural and peri-forest settings of Vellore, Chittoor and Tirupattur. This correlates with the seroprevalence data from northern Tamil Nadu, which showed a significant association with people residing in rural and peri-forested areas (19). This also explains the higher incidence observed among children and housewives, who are more likely to be exposed to peridomestic tick habitats influenced by climatic and ecological conditions (15). A prospective cohort-based study is required to determine the actual risk factors contributing to spotted fever occurrence in and around Vellore.

The higher case detection from areas close to the hospital may be due to improved access to specialised care and diagnostic testing. An increase in case testing, including both serology and molecular, was noticed from 2019. This was due to better awareness among the clinicians as a prospective study was conducted on spotted fever was conducted at our centre (1) . Similar observations have been reported following a surveillance study in the US (20) and with the inclusion of ELISA and PCR in Brazil (21).

The seasonality of spotted fever group rickettsioses demonstrates marked regional variation. In our study, the monthly trend aligns well with vector ecology. We observed a clear seasonal peak in January (IRR: 22.8, 95% CI (10.02 – 51.76), p <0.001), typically increasing from September to March, after which the number of cases declined. This pattern can be attributed to environmental factors such as sustained humidity, moderate temperatures, and increased vegetation, which enhance tick survival, reproduction, and host-seeking activity, ultimately leading to increased transmission during the cooler months (17). This retrospective analysis confirms the seasonal variation in spotted fever occurrence in and around Vellore. This cooler month surge in cases apparently contrasts with seasonality in European countries like Spain, Portugal and Italy (13,15,16). This is because the ambient temperature in these European countries matches the temperature in Southern India during the cooler months.

This study has limitations, as it is a single-centre study and does not include meteorological data. Therefore, the findings may not fully represent the broader epidemiology of spotted fever in and around Vellore.

## Conclusion

This study demonstrates the spatio-temporal pattern and seasonality of this rickettsial disease. This is characterised by clustering of cases in specific geographic areas in and around Vellore during the cooler months, with children being commonly affected.

## Data Availability

All data produced in the present study are available upon reasonable request to the authors

