## Supplementary data for "Spatio-temporal analysis of spotted fever cases reported to a tertiary care hospital in Southern India"

### Supplementary Tables & Figures

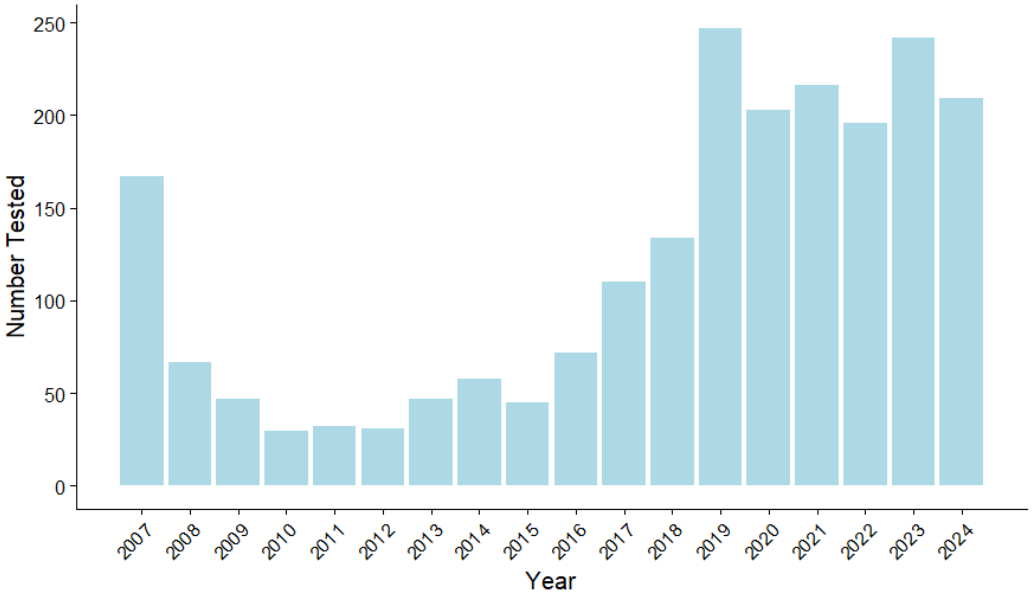


**Suppl Fig. 1: Number of SF cases tested annually**

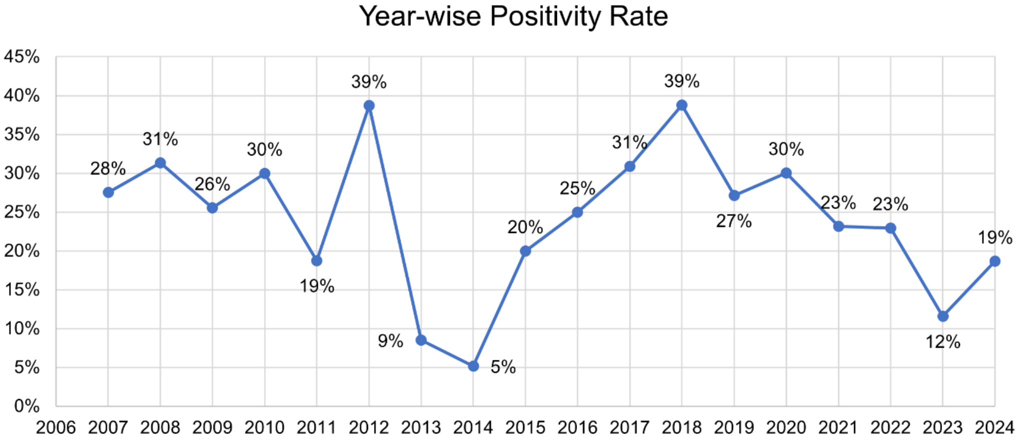


**Suppl Fig. 2: Year-wise SF positivity rate**

Suppl Table 1. Spatial distribution of SF Cases during the study period

| **Districts** | **2007-2012** | **2013-2018** | **2019-2024** | **Total** |
|---|---|---|---|---|
| Vellore | 30 | 48 | 98 | 176 |
| Chittoor | 45 | 36 | 90 | 171 |
| Tirupattur | 6 | 16 | 33 | 55 |
| Ranipet | 0 | 5 | 15 | 20 |
| Tiruvannamalai | 5 | 2 | 12 | 19 |
| **Total** | **86** | **107** | **248** | **441** |

Suppl Table. 2. Taluk-wise SF cases reporting in the 5 districts (2007-2024)

| District | Taluk | Number of cases |
|---|---|---|
| Vellore | Vellore | 65 |
| | Gudiyattam | 51 |
| | Katpadi | 40 |
| | Vaniyambadi | 8 |
| | Anaikkattu | 6 |
| | Pernambut | 6 |
| Chittoor | Chittoor | 44 |
| | Madanapalle | 17 |
| | Palmaner | 17 |
| | Thavanampalle | 11 |
| | Gangadharanellore | 7 |
| | Pileru | 7 |
| | Tirupati Urban | 7 |
| | Yadamari | 7 |
| | Irala | 5 |
| | Peddapanjani | 5 |
| | Tirupati Rural | 5 |
| | Bangarupalem | 4 |
| | Puthalapattu | 4 |
| | Chinnagottigallu | 3 |
| | Gangavaram | 3 |
| | Gudipala | 3 |
| | Pungunuru | 3 |
| | Srirangarajapuram | 3 |
| | Vayalpad | 3 |
| | Chandragiri | 2 |
| | Kalikiri | 2 |
| | Pakala | 2 |
| | Penumur | 2 |
| | Pulicherla | 2 |
| | Birangi Kothakota | 1 |
| | Kalakada | 1 |
| | Srikalahasti | 1 |
| Tiruppattur | Tiruppattur | 29 |
| | Ambur | 11 |
| | Vaniyambadi | 8 |
| | Natrampalli | 7 |
| Ranipet | Walajapet | 12 |
| | Arcot | 5 |
| | Arakkonam | 3 |
| Tiruvannamalai | Polur | 6 |
| | Arani | 6 |
| | Tiruvannamalai | 3 |
| | Vandavasi | 3 |
| | Kilpennathur | 1 |

Taluks with no case reported are not mentioned in the above table.
